# The breath volatilome is shaped by the gut microbiota

**DOI:** 10.1101/2024.08.02.24311413

**Authors:** Ariel J. Hernandez-Leyva, Amalia Z. Berna, Yang Liu, Anne L. Rosen, Michael A. Lint, Samantha A. Whiteside, Natalia Jaeger, Ryan T. McDonough, Nikhilesh Joardar, Jesús Santiago-Borges, Christopher P. Tomera, Wentai Luo, Audrey R. Odom John, Andrew L. Kau

## Abstract

The gut microbiota is widely implicated in host health and disease, inspiring translational efforts to implement our growing body of knowledge in clinical settings. However, the need to characterize gut microbiota by its genomic content limits the feasibility of rapid, point-of-care diagnostics. The microbiota produces a diverse array of xenobiotic metabolites that disseminate into tissues, including volatile organic compounds (VOCs) that may be excreted in breath. We hypothesize that breath contains gut microbe-derived VOCs that inform the composition and metabolic state of the microbiota. To explore this idea, we compared the breath volatilome and fecal gut microbiomes of 27 healthy children and found that breath VOC composition is correlated with gut microbiomes. To experimentally interrogate this finding, we devised a method for capturing exhaled breath from gnotobiotic mice. Breath volatiles are then profiled by gas-chromatography mass-spectrometry (GC-MS). Using this novel methodology, we found that the murine breath profile is markedly shaped by the composition of the gut microbiota. We also find that VOCs produced by gut microbes in pure culture can be identified *in vivo* in the breath of mice monocolonized with the same bacteria. Altogether, our studies identify microbe-derived VOCs excreted in breath and support a mechanism by which gut bacterial metabolism directly contributes to the mammalian breath VOC profiles.

## INTRODUCTION

Medical decision-making often relies on measurement of biomarkers that are associated with a disease state and provide prognostic or diagnostic information. While many clinical biomarkers require painful or invasive samples, there is increasing interest in the development of non-invasive clinical diagnostics. One promising avenue for biomarker identification is in exhaled breath, which is readily obtained and comprised of diverse compounds whose abundances are linked to human health (*1, 2*). Studies interrogating breath for biomarker discovery often focus on volatile organic compounds (VOCs): aromatic or low molecular weight molecules with high vapor pressure. Specific VOCs have been linked to respiratory diseases (*2–4*), as well as other diseases throughout the body (*5–8*). While some disease-associated VOCs reflect exposures to factors linked to pathogenesis or underlying changes in host metabolism and inflammation, the origin of many potential breath VOCs biomarkers is unclear.

In addition to compounds produced endogenously by the body, many VOCs emitted by humans are derived from the metabolism of host-colonizing microbes (*9–11*). Changes in breath VOC composition have been previously identified in subjects infected with tuberculosis or malaria (*12–16*), some of which may be produced by the pathogen and modulate microbial pathogenicity (*15*). Bacterially produced VOCs are diverse, can reflect the growth state of the bacterium, and are known to serve valuable functions in quorum sensing and immune modulation (*17–24*). Emitted VOCs from soil Actinomycetota can even serve as a conserved taxonomic signature, similar to the conserved regions of the 16S ribosome (*25*).

The gut microbiota, the sum of all prokaryotic microorganisms residing within the intestines of a living host, has been extensively implicated in human disease (*26*). Changes in the gut microbiota have been identified in association with disease locally in the intestines (*27*) and distally in extraintestinal tissues such as the lung (*28, 29*), liver (*30*), and brain (*31*). Gut microbes produce a diverse set of metabolites that disseminate throughout the body, even into anatomically protected tissues (*32, 33*). Many of the VOCs produced in the gut are subject to first-pass metabolism in the epithelium or liver, converting compounds like phenol to p-cresol(*34*) or trimethylamine to trimethylamine-N-oxide (*35–37*), which can be renally excreted. However, many of the volatiles present in exhaled breath including acetate, acetone, and ammonia are also major products of microbial metabolism (*38–40*). A catalog of the VOCs of exhaled breath that are influenced by the gut microbiota would be important for identifying reliable biomarkers of human disease in breath. Moreover, identifying to what degree changes in the breath volatilome reflect changes in the gut microbiota would open new avenues toward real-time monitoring of human gut health. Rapid, point-of-care assessment of the state of the gut microbiome could greatly improve clinical care, particularly in clinical settings such as necrotizing enterocolitis and serious bacterial infection in preterm infants (*41*), where prompt recognition could trigger early intervention.

Here, we test the hypothesis that gut microbial metabolism directly contributes to the breath VOC profile of mammalian hosts. Thus, host breath biomarkers may represent changes not only in host metabolism but also to the metabolism and composition of the gut microbiota. To address this idea, we interrogate the breath and gut microbiota of a cohort of 27 healthy children between the ages of 6 - 12 years and demonstrate that the composition of the gut microbiota helps shape the breath VOC profile of humans. To test this idea, we devise and validate a gnotobiotic mouse model for breath collection. Using this model, we study the influence of individual gut microbes on breath by comparing the volatile emissions of common gut microbes grown under anaerobic conditions to the breath of mice colonized with a single bacterial strain. Together, these experiments show that the gut microbiota helps shape the host breath volatile profile and establishes models to further interrogate the relationship of the gut microbiota with breath biomarkers.

## RESULTS

### The breath volatilome of healthy children reflect the composition of the gut microbiota

We collected stool and breath samples from a study of 27 healthy children (ages 6 - 12 years) to determine whether there was a correlation between the composition of the gut microbiome and the VOC composition (volatilome) of exhaled breath. No children received antibiotics within a month of study enrollment and tobacco exposure was low across the cohort **(Table S1)**.

We first characterized the human stool microbiome by whole metagenomic sequencing (**Table S2**). The overall phyla-level composition of the gut metagenome was consistent with published reports of the human metagenome, and was dominated by Bacillota, Bacteriodota, and Actinomycetota with smaller but significant contributions from Pseudomonadota, Fusobacteriota, and Verrucomicrobiota (*42*) (**Fig. S1A**). We next compared demographic features of our cohort to the UniFrac distances derived from metaphlan taxonomic assignments to identify factors shaping the gut microbiota composition and assess for potential confounding variables. Consistent with prior reports, we identified that pet ownership influenced the overall taxonomic composition of the gut metagenome that could not be attributed to differences in beta dispersion (**Fig. S1B-C),** PERMANOVA R2 = 0.060, *p-value* = 0.038, PERMDISP *p-value* = 0.5112]. While underpowered for such analysis, other factors including sex, BMI, and age did not significantly contribute to the observed variance in gut microbiota composition.

Next, we determined the composition of the breath volatilome for each participant. For each patient, breath VOCs were captured onto sorbent material and subsequently released by thermal desorption for analysis by gas chromatography and time-of-flight mass spectrometry (GC-qToFMS). We identified a total of 2734 features across all subjects, which represent the unique combination of a specific value of mass-to-charge ratio (m/z) and GC retention time (RT) of a molecular ion. We also assessed the potential influence of demographic factors on the overall composition of exhaled breath volatiles, but found no significant relationship with age, sex, BMI, race, ethnicity, or pet ownership by PERMANOVA (**Fig. S1D-E**).

To determine whether the composition of the gut metagenome correlated with the VOC profile of exhaled breath we performed a Procrustes rotation with PROTEST (*43*). Procrustean rotations provide a statistical method to ask whether the interindividual differences between breath volatilomes in these children correlate with the interindividual differences amongst their gut microbiota. We first compared the distribution of exhaled breath estimated using Manhattan distances to the taxonomic composition of the gut metagenome estimated using UniFrac distances (*44*) and identified a significant correlation (**Fig. 1A**, PROTEST Correlation: 0.74, p-value = 0.023). Furthermore, the exhaled breath profile correlated with both the composition of uniref90 gene clusters (**Fig. 1B**, PROTEST Correlation: 0.79, p-value = 0.011) as well as functional pathways (**Fig. 1C**, PROTEST Correlation: 0.71, p-value 0.03) inferred by humann3. Together these data suggest a link between the composition and function of the gut microbiota and the breath volatilome.

**Fig. 1:**
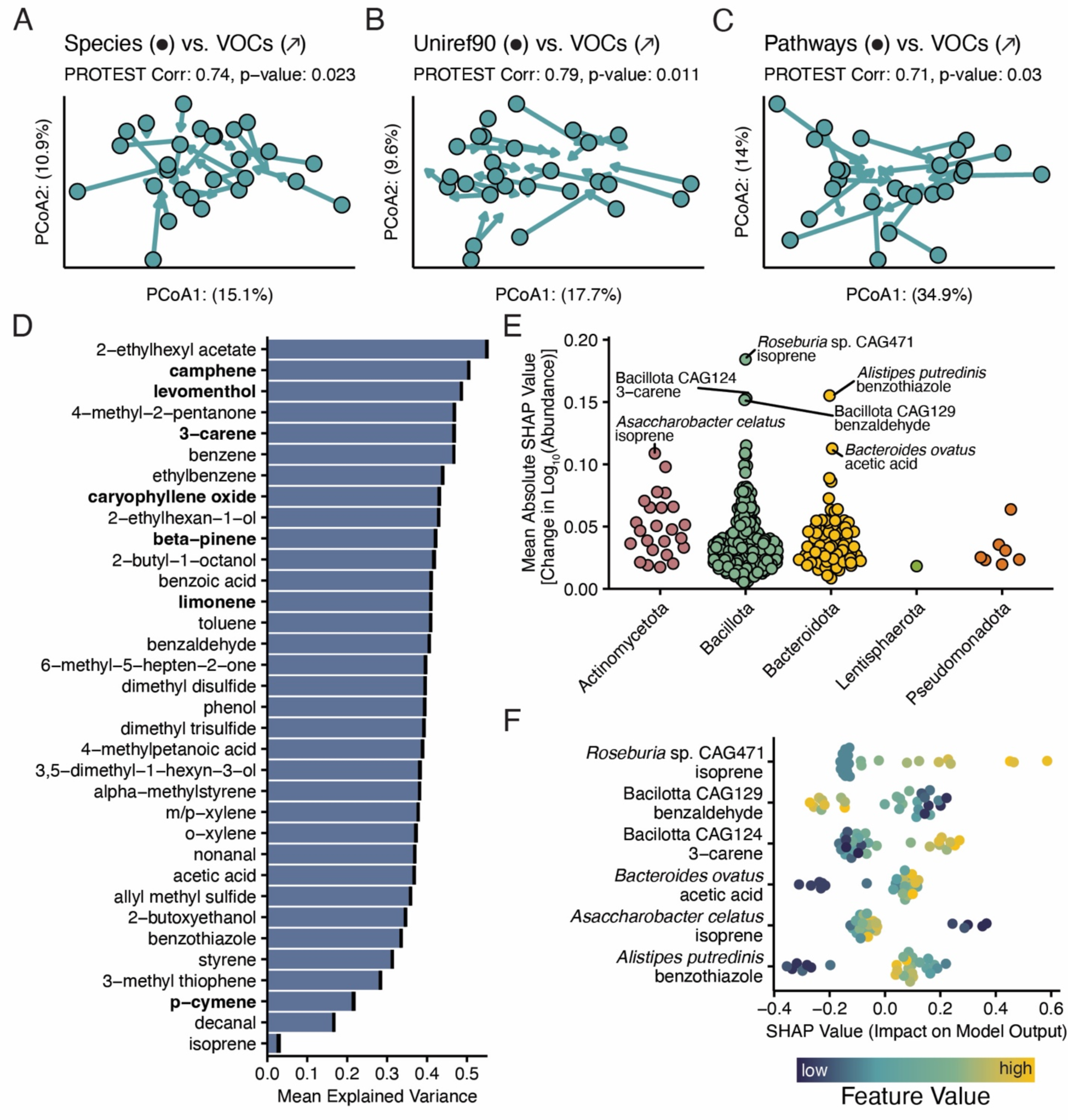
The pediatric gut microbiome influences the host breath volatilome. A) Procrustean rotation comparing the UniFrac distances describing subject microbiome taxonomic composition (points) against the Manhattan distances comparing subject breath volatilome (arrows). B) Procrustean rotation comparing the Manhattan distances describing the abundances of subject gut uniref90 gene clusters (points) against the Manhattan distances comparing subject breath volatilome (arrows). C) Procrustean rotation comparing the Manhattan distances describing subject gut metagenomic pathways (points) against the Manhattan distances comparing subject breath volatilome (arrows). D) The mean explained variances (R^2^) of individual random forest models predicting the abundance of a particular VOC given the taxonomic composition of the gut metagenome. Terpenes are bolded. E) Plot of the importance of individual taxa across the models, estimated as the mean of the absolute value of the SHAP scores for that taxon in its respective model. Individual points represent the Importance of a taxon in one of the models. F) Sample SHAP scores for the top seven most important model/taxa pairings. Feature values were ranked to show trends against the SHAP score.

Since identifying robust associations between thousands of GC-MS features in breath and hundreds of taxa in the gut microbiota would pose statistical challenges, we focused on characterizing a subset of VOCs identified from the breath literature (*1*), which are reproducibly found in the breath of humans (**Table S3**). To determine if specific bacterial taxa contributed to the presence of these breath metabolites, we built random forest models with feature selection to predict the abundance of each VOC from the relative abundances of the 302 gut taxa found in our cohort. Importantly, for many of the selected VOCs, substantial variance (up to 40%) in breath abundance is explained by changes in the composition of the gut microbiota (**Fig. 1D**). Moreover, the gut microbiota compositional data explained 20% or more of the variance of 35 of the 37 VOCs on our curated list of breath VOCs. Notably, many of the most microbially influenced breath VOCs (e.g., camphene, 3-carene, beta-pinene, limonene, and p-cymene; bolded) belong to a class of compounds known as terpenes. Terpenes are a family of highly diverse natural products produced by plants and microbes (*45, 46*), but not mammals, and breath terpene levels have been associated with a number of disease conditions, including malaria, fungal infections, and liver disease (*15, 47–49*).

We also sought to identify whether specific taxa were associated with specific breath VOCs, by calculating the mean absolute value of the Shapley addictive explanations (SHAP values, **Fig. 1E**). We visualized the SHAP value per sample to estimate the directionality of the relationship between the VOC and taxa (**Fig. 1F)**. This analysis revealed particularly strong associations between several bacteria and common breath VOCs. For example, there was a positive relationship between the *Bacillota* bacterium CAG124 and the terpene 3-carene. This analysis also identified interactions between taxa and their known metabolic products, such as between *Bacteroides ovatus*, a well-known producer of short chain fatty acids (SCFA), and the SCFA acetic acid (*50*).

### Establishing a model for studying the influence of the gut microbiome on exhaled breath using gnotobiotic mice

While our human study suggests an association between breath VOCs and gut bacteria, demonstrating a causal relationship between the two requires an animal model in which we have robust control of the gut microbiome. To develop further insights into the gut microbial origins of breath VOCs, we devised a method to collect the exhaled breath of gnotobiotic mice using the SCIREQ Flexivent FX1 murine ventilator (*51, 52*). This small animal ventilator uses a unidirectional airflow ideal for the capture of exhaled breath with minimal contamination. As an additional benefit, this system typically requires ventilating mice through a cannula surgically implanted into the trachea, thus maximizing collection of alveolar air over VOCs arising from the oral cavity (**Fig. 2A**).

**Fig. 2:**
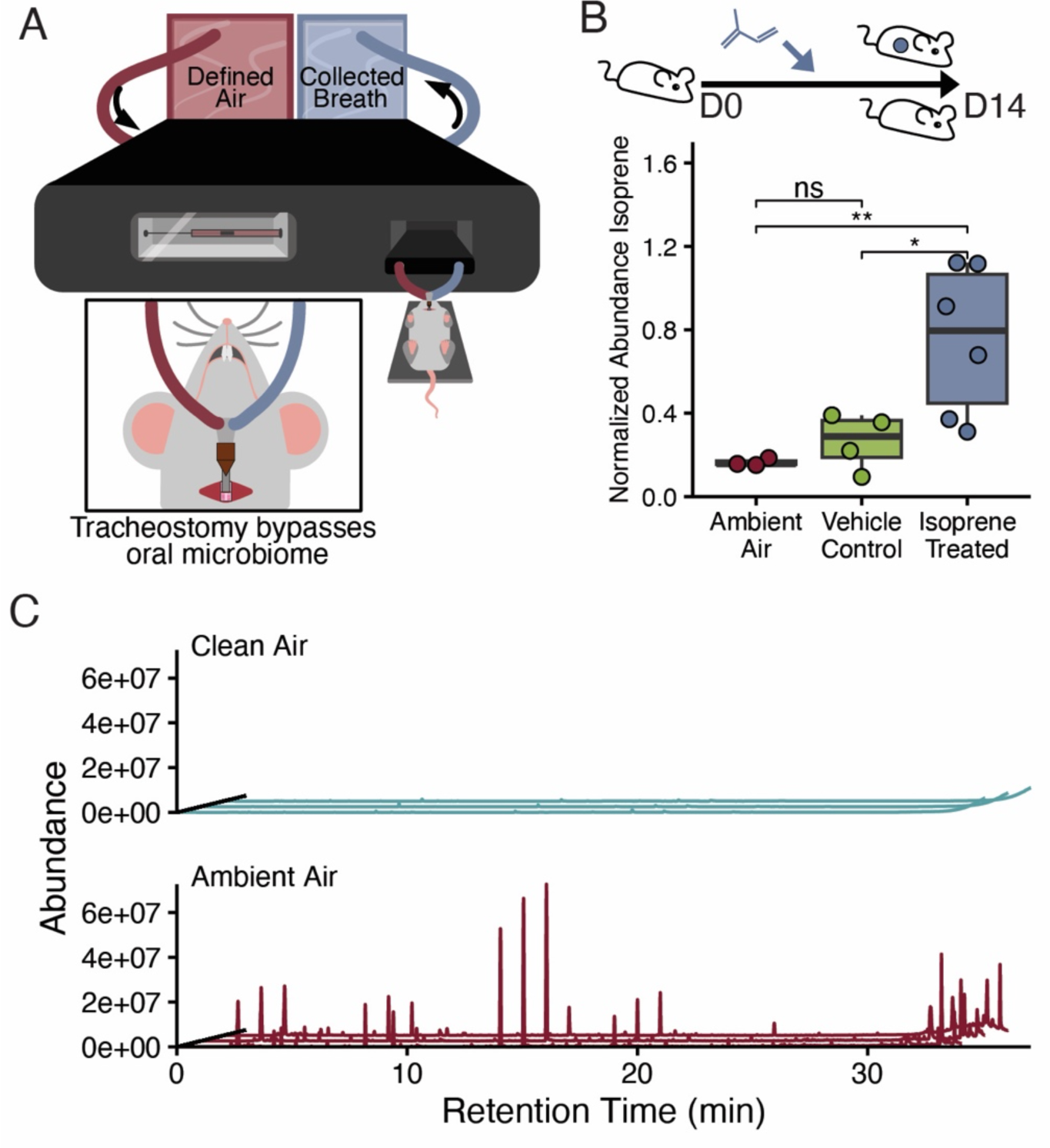
A murine ventilator can be used to collect murine exhaled breath. A) Overview of the mouse ventilator set-up to collect exhaled breath while providing defined air. B) Box plot representing the abundance of isoprene in the exhaled breath of mice receiving daily isoprene doses for 14 days, in exhaled breath of mice receiving concurrent daily vehicle controls, or in the ambient air alone. Between group comparisons were conducted using t-tests with FDR multiple hypothesis correction. C) Representative chromatograms depicting the signal measured from output air collected from the ventilator after attaching an airtight plastic loop instead of a mouse. Signals were characterized from air collected with defined input air or input air collected from an empty conference room.

To validate this method, we designed a proof-of-concept experiment in which we introduced an exogenous VOC, isoprene, that we could then measure in captured mouse breath. Isoprene is a common VOC in the exhaled breath of many mammals (*53*). Previous studies of pharmacokinetics of isoprene in mice and rats demonstrated that a relatively small fraction of endogenously produced isoprene is exhaled (*54*), with most of the isoprene undergoing further metabolic alterations. However, exogenous administration of isoprene can significantly increase exhaled isoprene in rats (*54*). To test if our breath collection protocol would capture isoprene, we intraperitoneally (IP) injected C57BL/6 mice with 1 mg of isoprene or vehicle alone once per day for fourteen days before collecting breath. Mice were not injected on the day of collection.

Since isoprene preparations were prepared in the same laboratory space as breath collection, we also collected ambient air controls to ensure measured isoprene levels were not a result of contamination with laboratory air. We quantified isoprene in the samples using GC-MS and confirmed the identity of isoprene using an analytical standard (see Methods). Our results showed increased abundance of isoprene following administration of isoprene, compared to vehicle alone (**Fig. 2B**). Additionally, a greater amount of isoprene was detected in the breath of mice receiving vehicle control compared to ambient air alone, indicative of endogenously produced isoprene.

After demonstrating reliable capture of murine breath VOCs, we further optimized our protocol by providing synthetic air to mice undergoing breath collection to minimize the introduction of external contaminants. We found that the use of synthetic air substantially reduced background contamination from indoor volatiles (**Fig. 2C**). Additionally, mouse-free input air controls were also performed before and after all breath collection events to enable the identification and elimination of remaining contaminants. Together, these results affirmed that our method for breath collection could capture and quantify differences in VOCs in the exhaled breath of mice, while minimizing the impact of exogenous contaminants.

### The exhaled breath VOC profile of mice is influenced by the composition of the gut microbiome

Using our optimized breath collection protocol, we next sought to test the effect of the gut microbiota on exhaled breath of mice. To perform these experiments, we took advantage of the observation that mice sourced from different facilities have distinct gut microbiota profiles. Thus, conventionally-raised male and female mice were obtained from two separate vendors, The Jackson Laboratory and Taconic Biosciences, typically associated with distinct gut microbiota compositions (*55, 56*). After one week, breath was collected from each mouse and fecal specimens were collected for characterization of the gut microbiome. Next, to test whether breath VOCs could be transferred by gut microbiota transplant, cecal contents from conventional mouse groups were homogenized and used to colonize sex-matched germ-free mice by oral gavage. One week after colonization, we again collected breath and fecal samples from these “conventionalized” mice. Exhaled breath from all mice was characterized using GCxGC-MS and fecal specimens were analyzed by whole metagenomic sequencing (**Fig. 3; Table S2**). At all breath collection events, sex-matched germ-free mouse breath was collected as a control. Importantly, all mice, including the conventional mice, were housed under identical conditions in the same gnotobiotic facility and fed the same diet for at least one week prior to breath collection (see Methods).

**Fig. 3:**
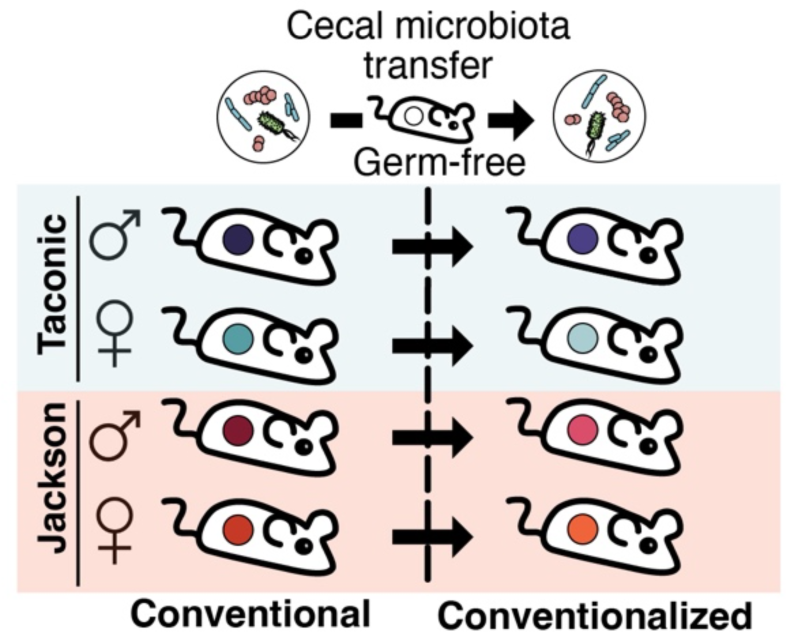
Overview of a gnotobiotic animal model for testing the contribution of the gut microbiota to the breath volatilome. A) The exhaled breath of conventionally raised male and female mice sourced from either Taconic Biosciences or the Jackson Laboratory was collected and characterized. The cecal contents of the conventionally-raised mice from the same vendor and of the same sex were pooled and used to colonize sex-matched gnotobiotic recipients, yielding four more microbiome groups: male gnotobiotic mice colonized with cecal contents from male Jackson Lab mice, female gnotobiotic mice colonized with cecal contents from female Jackson Lab mice, male gnotobiotic mice colonized with cecal contents from male Taconic Biosciences mice, female gnotobiotic mice colonized with cecal contents from female Taconic Biosciences mice. Breath was collected and characterized from these mice one week after colonization. Breath was collected from germ-free control mice at all timepoints.

We first assessed the gut composition of the donor and recipient mice. As expected, transfer of microbes between mice was efficient (**Fig. S2A**), with engraftment scores of greater than 98%. We also used principal coordinates analysis on the UniFrac distances to understand how the mouse microbiota were related to one another (**Fig. S2B**). As expected, recipient mice clustered near their respective donors; however, groups of mice were significantly different from one another (PERMANOVA R^2^: 0.173, p-value < 0.0002) indicating that transplantation did have some effect on the gut microbiota. The recipient mice possess microbiota largely similar to that of their donors; however, there are notable differences in the relative abundances of many taxa (**Fig. S2C**). For example, *Akkermansia muciniphila*, present in three of the four donor microbiota, is found at a greater relative abundance in the conventionalized recipients than in their respective donors.

We next analyzed the composition of exhaled breath from conventional and conventionalized mice as well as germ-free controls by GCxGC-MS. We visualized processed and filtered breath metabolite data using a principal coordinates analysis on Manhattan distances between subjects (**Fig. S3A**). We determined that the individual microbiome had a substantial impact on breath volatile composition, finding that the individual microbiome group explained more than 20% of the variation in the breath profiles of a given mouse (PERMANOVA R2: 0.222, p-value = 0.0024). As previously identified in humans (**Fig. 1A-C**), we found that the interindividual variation in the breath profile correlated with that of the gut microbiota. Specifically, the taxonomic profile (**Fig. 4A**, PROTEST Correlation 0.75, p-value = 0.0047) and uniref90 cluster compositions (**Fig. 4B**, PROTEST Correlation 0.68, p-value: 0.006) correlated with the breath VOC profile. While the pathway composition of the gut microbiota did strongly correlate, PROTEST did not find a significant association (**Fig. 4C**, PROTEST Correlation 0.68, p-value: 0.054).

**Fig. 4:**
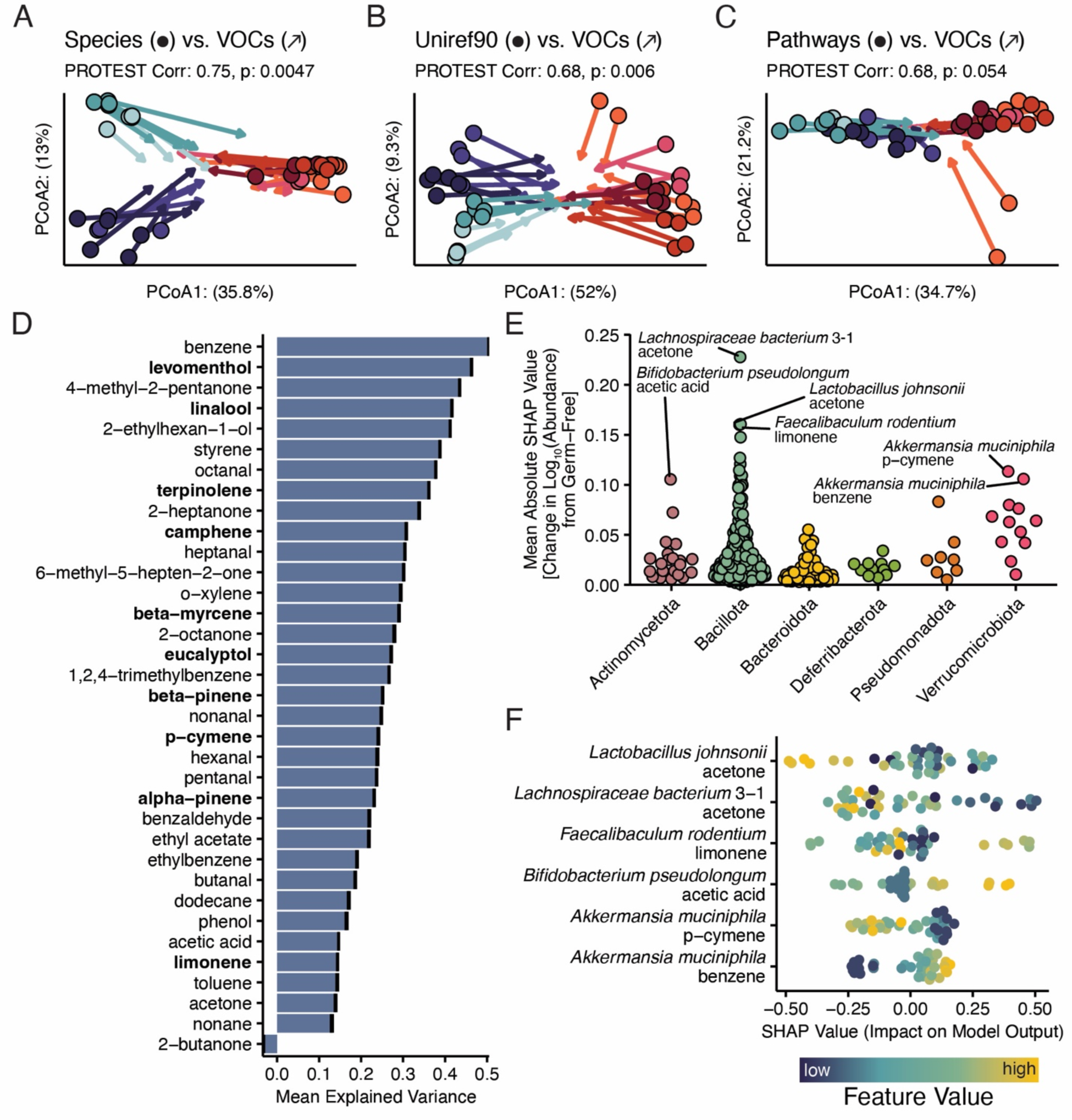
The gut microbiota influences the host mouse exhaled breath volatilome. A) Procrustean rotation comparing the UniFrac distances describing mouse microbiome taxonomic composition (points) against the Manhattan distances comparing subject breath volatilome (arrows). B) Procrustean rotation comparing the Manhattan distances describing the abundances of mouse gut uniref90 gene clusters (points) against the Manhattan distances comparing mouse breath volatilome (arrows). C) Procrustean rotation comparing the Manhattan distances describing mouse gut metagenomic pathways (points) against the Manhattan distances comparing mouse breath volatilome (arrows). D) The mean explained variances (R^2^) of individual random forest models predicting the abundance of a particular VOC given the taxonomic composition of the gut metagenome of mice. Terpenes are bolded. E) Plot of the importance of individual taxa across the models, estimated as the mean of the absolute value of the SHAP scores for those taxa in its respective model. Individual points represent the Importance of a taxa in one of the models. F) Sample SHAP scores for the top seven most important model/taxa pairings. Feature values were ranked to show trends against the SHAP score.

Using the curated list of common breath VOCs described above (**Table S3**), we extracted individual features representing these VOCs in the breath of mice to further explore specific VOC-microbe associations. To control for day-to-day variance in contaminating VOCs, the log2 fold change for each VOC between colonized mouse breath and germ–free controls collected on the same day was calculated. We found that 22 of 35 breath VOCs varied significantly between experimental groups (**Fig. S3B**, including benzene and ethyl acetate, indicating that the microbiota and conventionalization impacted the abundance of these compounds in breath. We also used random forest modeling with feature selection (see also **Fig. 1E**) to predict the change from germ-free breath of each VOC from the relative abundances of the gut taxa. These models showed that the gut microbiota composition explained at least a part of the variance for 34/35 of the interrogated compounds (**Fig. 4D**). Of the 5 breath VOCs best explained by gut microbiota composition, all had greater than a 40% mean variance explained. Several compounds, such as 4-methyl-2-pentanone, benzene, and levomenthol, shared a relatively high variance explained by the gut microbiota in both human (**Fig. 1D**) and mouse breath. As before, we estimated the importance of each taxa to each model using the mean absolute SHAP values (**Fig. 4E**). We also visualized the relationship between the top few most important taxa-VOC pairings by plotting the sample SHAP values (**Fig. 4F)**. Among the interesting VOC-microbe relationships observed here, the negative association of both *Lachnospiraceae* bacterium 3-1 and *Lactobacillus johnsonii* with acetone is particularly notable. Additionally, we noted that *A. muciniphila* colonization corresponded with increases of benzene and reduced amounts of the terpene p-cymene.

### Human gut anaerobic bacteria produce identifiable VOCs under anaerobic conditions

While our analyses in gnotobiotic animals support the idea that the gut microbiota influences the breath VOC profile of the host, these experiments used complex consortia of gut bacteria, which limits the ability to assess the contributions of any single member of the gut community to breath VOC profiles. To evaluate whether individual gut bacteria produce VOCs measurable in breath, we sought to characterize the VOC profile of individual bacteria grown in pure culture. We developed a method to collect culture headspace VOCs from gut bacterial cultures grown under anaerobic conditions in a rich media supplemented with mucin, to approximate the metabolism of bacteria in the gut environment (**Fig. 5A**). To demonstrate that our culture headspace collection approach was capable of discerning VOCs produced by gut bacteria under anaerobic conditions, we focused on production of a well-characterized volatile metabolite (indole) by *B. thetaiotaomicron*. Indole is produced from tryptophan by the tryptophanase enzyme encoded by the gene *tnaA* in *B. thetaiotaomicron* (*57*). Comparison of the levels of indole in anaerobic culture headspace between wild-type *B. thetaiotaomicron* and *tnaA* deficient isogenic *B. thetaiotaomicron* showed that indole was enriched only in the headspace of wild-type bacteria compared to sterile media controls or the *tnaA* deficient strain (**Fig. 5B**). We next collected culture headspace of three additional bacteria, *A. muciniphila*, *Escherichia coli*, and *Bifidobacterium longum*, and after analyzing headspace samples using GC-MS we compared the VOC profile of culture headspace for these strains against sterile media control (**Fig. 5C**). We found that each bacteria produced a headspace VOC profile that was distinct from media and one another, suggesting that taxonomically distinct bacteria produce unique VOC profiles.

**Fig. 5:**
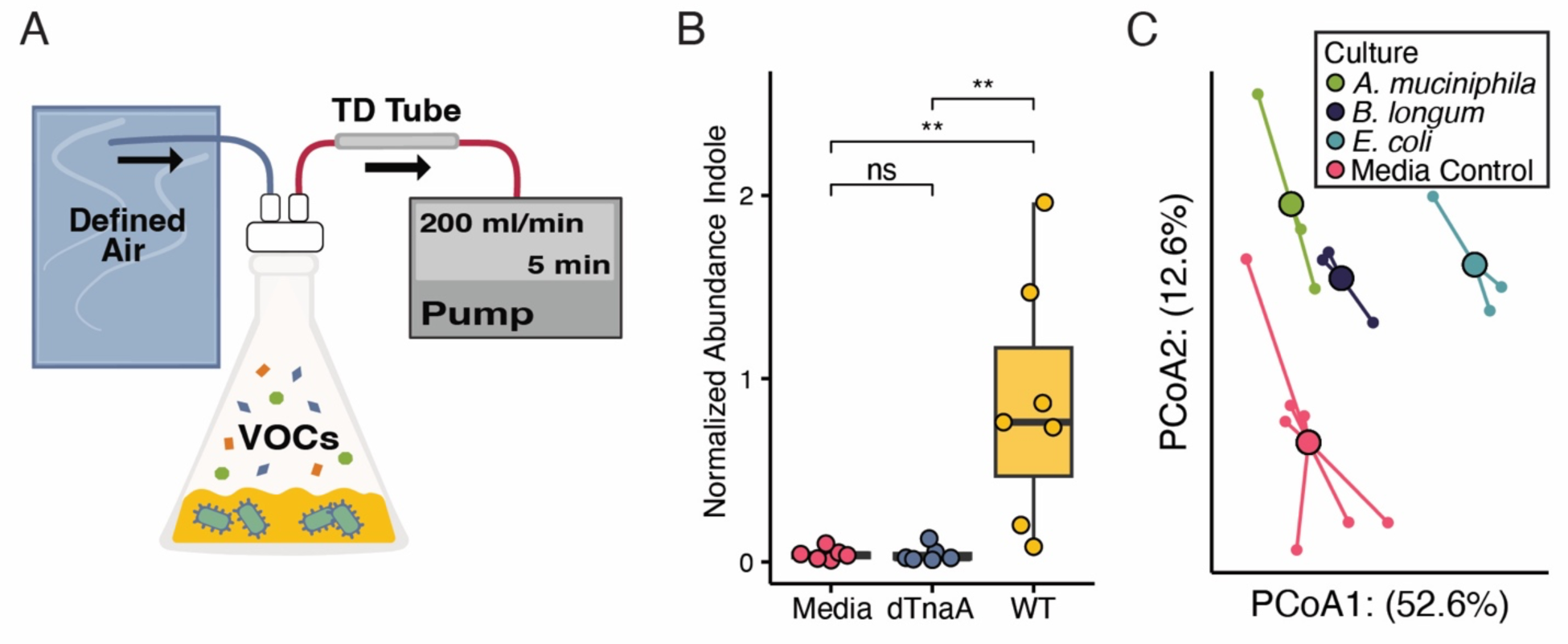
Collection of microbial derived VOCs from anaerobic bacteria. A) Cartoon depicting the collection of anaerobic culture headspace by drawing defined anaerobic air through the headspace of confluent culture and across a thermal desorption tube. B) Box-plot depicting the abundance of indole in the headspace of confluent culture of WT *B. thetaiotaomicron*, confluent culture of *B. thetaiotaomicron* deficient in tryptophanase (ΔtnaA), and sterile media controls. Between group comparisons were conducted using Mann-Whitney tests with Benjamini-Hochberg multiple hypothesis correction. C) Principal coordinates analysis depicting Manhattan distances between the volatilomes of the culture headspaces of *A. muciniphila, B. longum*, *E. coli*, and sterile media controls.

To test the idea that some breath VOCs arise directly from gut bacterial metabolism, we colonized gnotobiotic mice with one of five common gut commensals including *A. muciniphila*, *B. thetaiotaomicron*, *Collinsella aerofaciens*, *E. coli*, and *Ruminococcus torques* and sterile media controls. Additionally, we collected headspace VOCs from anaerobic cultures of each bacterium. After two weeks of colonization, we collected exhaled breath from each group of monocolonized mice as well as germ-free controls. All culture headspace and mice breath samples were then characterized using a GCxGC-MS. We then employed a feature selection algorithm (ChromCompare+) to identify differentially abundant VOCs from both anaerobic culture and sterile media headspace or monocolonized and germ-free mouse breath. We employed a bidirectional approach to screening so that when one compound was identified as differentially abundant in one sample type, the same compound was also screened in the profiles from the other sample type. In addition to identifying VOCs associated with a single bacterial species like ethyl acetate and *E. coli*, this approach also identified several VOCs associated with multiple species in culture such as 2-nonanone which was associated with *C. aerofaciens*, *E. coli*, and *R. torques* (**Fig. 6A; Table S4**). Similarly, we found multiple VOCs that were enriched in gnotobiotic mice monocolonized with different bacteria, such as m- and o-xylene and toluene (**Fig. 6B; Table S4**). These are compounds that have been observed in human breath, but thought to be associated with exposure to pollutants and smoking (*1*).

**Fig. 6:**
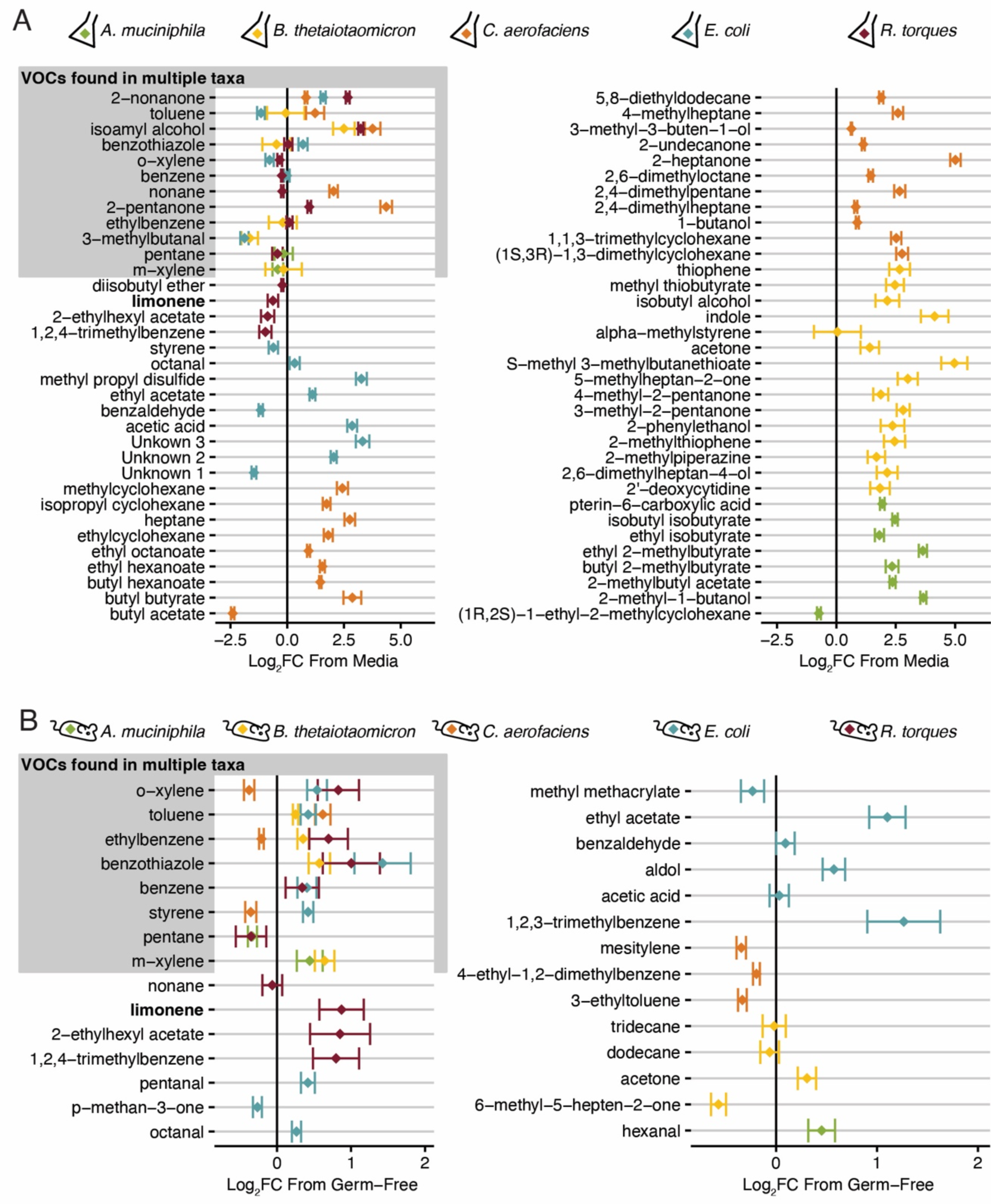
Overview of discriminatory microbially-associated VOCs in anaerobic culture headspace and the exhaled breath of monocolonized mice. A) Forest plot denoting the 95%-confidence interval around the mean log2 fold difference between a VOC in the anaerobic culture headspace of a given organism and respective media controls. Confidence interval was estimated using the pooled variance of both culture and media abundances of VOCs. B) Forest plot denoting the 95%-confidence interval around the mean log2 fold difference between a VOC in the breath of mice colonized with a given organism and the breath of germ-free controls. Confidence interval was estimated using the pooled variance of both colonized and germ-free mouse breath abundances of VOCs. Terpenes are bolded.

Finally, we queried whether there were individual compounds that were enriched in both in the culture headspace and breath of mice monocolonized with the same organism. This comparison revealed that culture headspace and monocolonized mouse breath associated with *E. coli* was enriched for benzothiazole, ethyl acetate, and octanal (**Fig. 7**). Acetone and toluene were likewise enriched in both the culture headspace and monocolonized mouse breath associated with *B. thetaiotaomicron* and *C. aerofaciens*, respectively. In contrast, pentane was reduced in both the culture headspace and breath of mice colonized with *R. torques*. Notably, there were several compounds that were both enriched in colonized mouse breath, but depleted compared to media, including 2-ethylhexyl acetate, benzene, and styrene.

**Fig. 7:**
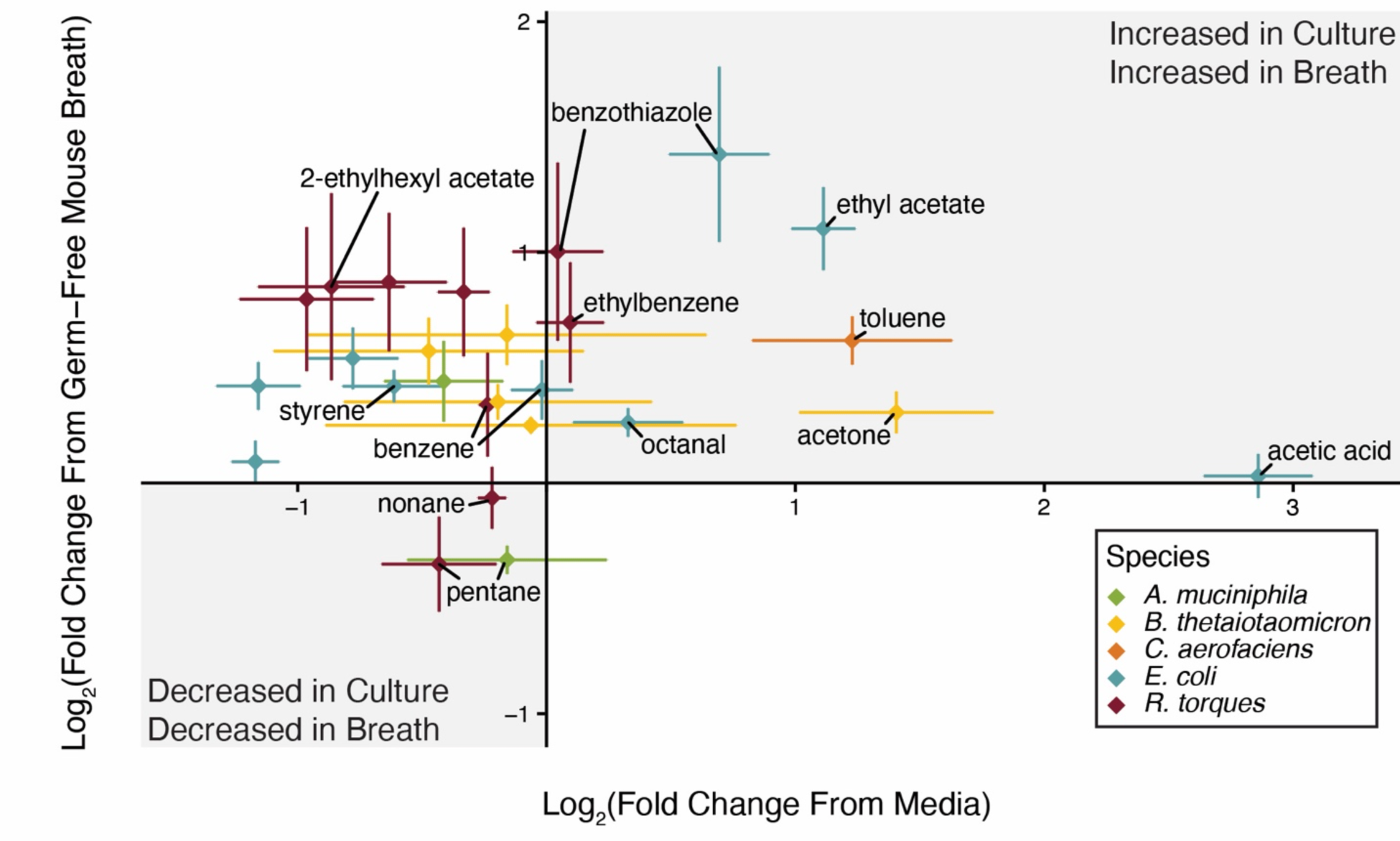
Comparing the volatilome of anaerobic culture headspace to the exhaled breath of monocolonized mice identifies potential gut microbiota-derived breath VOCs. Plot comparing the means and 95% confidence-intervals of the log2 fold difference in VOC abundances between culture headspace or colonized mouse breath and media headspace or germ-free mouse breath, respectively. Confidence intervals were estimated using the pooled variance of both colonized/culture samples and germ-free/media controls.

## DISCUSSION

In this study, we applied a multi-omic analysis of a unique cohort of human subjects and a newly developed experimental mouse model to evaluate breath volatile profiles and microbiome composition, to determine the contribution of gut microbes to the breath volatilome. We find that the breath volatilome is correlated with the composition of the gut microbiota. In addition, the variance of several common human breath VOCs can be explained, at least in part, by the changes in the abundance and functions of gut microbes. Some of these compounds are expected products of microbial fermentation in the gut, such as acetic acid, a short chain fatty acid commonly produced by *Bacteroides* species (*50*). Previous studies examining crosstalk between the breath and gut have focused primarily on the results of an intervention in human subjects, such as changes in diet or the difference between health and disease (*7, 58, 59*). However, these foundational studies offer limited control over many of the factors that affect the breath volatilome composition, such as diet and environmental exposures (*60*). Our clinical study provides new evidence linking variation in the gut microbiota to changes in the breath volatilome of the host, in both mouse and humans.

To facilitate discovery of interactions between specific taxa and volatiles in breath, we developed a gnotobiotic mouse model to control for host and environmental factors impacting the breath volatilome to identify specific microbe-associated VOCs in breath. We show through our conventionalization experiments that the presence and composition of the gut microbiota of mice are also linked to its breath volatilome. Furthermore, by identifying microbially produced VOCs under anaerobic conditions, we establish an approach for finding biologically meaningful microbially derived VOCs in the breath volatilome. For example, ethyl acetate is elevated in the breath of patients recovering from extensive antibiotic use (*61*). In animal models, early microbial successors of a gut recovering from antibiotics often include Enterobacteriaceae such as *E. coli*, suggesting that exhaled breath compounds like ethyl acetate may represent a clinically useful biomarker that reflects recovery of the gut microbiome of patients.

Gut microbes may influence breath VOCs through multiple mechanisms. ***First***, it is likely that at least some VOCs are produced by gut bacteria and are absorbed and distributed to the lung alveoli, where they partition into breath exhalate. This model is supported by the results of our monocolonization studies, which demonstrated that several VOCs were found both in the culture headspace of a bacteria and exhaled breath of a mouse colonized with the same microbe. This model of microbially derived VOC production appears the most likely mechanism to explain increases in breath toluene in *C. aerofaciens*-colonized mice and ethyl acetate in *E. coli*-colonized mice. We further suspect that the gut microbiota as a whole may influence levels of major fermentation products, such as acetate, through this mechanism. ***Second***, compounds produced in the gut by microbes may undergo biotransformation by the mammalian host (e.g. methanethiol) (*62*), either in the epithelial cells or liver, prior to breath exhalation. ***Third***, the presence of the gut microbiota could impact the expression of hormones or metabolic enzymes in the host, such as insulin (*63*) or the cytochrome p450 enzymes in the liver (*64*), which could, in turn, alter the VOC profile of exhaled breath (*65–69*). ***Fourth***, many breath VOCs are thought to be of dietary origin, such as terpenes like eucalyptol and limonene (*70–72*). The gut microbiota may influence host breath composition by regulating intestinal barrier permeability and thus bioavailability of dietary metabolites. However, as many gut bacterial species also express terpene synthases, and breath terpene concentrations are highly influenced by the gut microbiome, breath terpenes may reflect a combination of both dietary and microbially produced metabolites. It is likely that all these processes help to shape the overall breath volatilome highlighting the importance of a well-controlled experimental model in disentangling gut microbe-breath VOC interactions.

Our clinical study to investigate the gut microbiota and breath of children was designed as a pilot study, and we therefore focused our investigations on a select set of well-characterized breath VOCs. Future larger studies are needed to comprehensively evaluate the thousands of features present in human breath. In addition, longitudinal studies are required to address the plasticity of the relationship between breath VOCs and changes in the gut microbial community, for example, following antibiotic exposure. As we gain additional insights into the relationship between specific gut taxa and breath VOCs, breathomics may ultimately represent a powerful diagnostic tool to facilitate rapid, non-invasive interrogation of gut microbial health and disease.

## MATERIALS AND METHODS

### Experimental Design of Human Study

The Microbiome-Asthma Connection in Kids (MACK) study was designed to test whether the breath volatilome might be used as a biomarker for dysbiosis in the gut microbial communities of subjects with asthma. The study, initiated in 2018, aimed to collect stool samples and breath samples from children with and without asthma. Recruitment for MACK was prematurely closed in 2020 by the SARS-CoV-2 pandemic. Since the goal of this study was to establish a relationship between the gut microbiota composition and breath VOC profiles in healthy individuals, the asthmatic subjects were not included in this analysis. Thirty-three total subjects were enrolled in the healthy cohort, and 27 of these had matched stool and breath samples.

Subjects admitted to the healthy cohort were 6 - 12 years of age at the time of enrollment and had neither a history of chronic illness nor any recent acute illness. Subjects were excluded from the healthy cohort if they had a history of wheeze or shortness of breath within 1 year of recruitment or any history of asthma, allergic rhinitis, food allergy, or eczema. Subjects were also excluded if they received non-topical antibiotics or corticosteroids within 30 days of recruitment, a diagnosis of any serious medical condition including insulin-dependent diabetes mellitus, or had surgery affecting the sinuses, lungs, or gastrointestinal tract. Demographic information collected, including subject age, sex, race, and ethnicity, is summarized in **Table S1.**

MACK was approved by the Washington University Institutional Review Board (IRB#201810180). Written and informed consent documents were obtained from each subjects’ parents or guardians.

### Breath Collection

Breath collection was performed as previously described (*15, 73*). In brief, subjects refrained from brushing their teeth, eating, or drinking for 2 hours prior to sample collection. Subjects were allowed to rest in an isolated room for several minutes before breath collection. Subjects exhaled through a disposable cardboard mouthpiece connected to a chamber. The chamber was then attached using tubing to a 3 L SamplePro FlexFilm sample bag (SKC Inc, Cat# 237-03). Subjects were coached to take a few deep breaths, place the cardboard tube between the lips, and exhale completely. Exhalation was completed 2 - 3 times as necessary to collect a minimum of 1 L of exhaled breath. Contents of human breath were pulled through CAMSCO inert stainless steel thermal desorption tubes (CAMSCO Cat# SIU60520-60-B) with an electric air pump at a rate of 200 mL/min for 5 min. Thermal desorption (TD) tubes were stored at 4° C in a sealed plastic container for no more than 2 weeks before analysis using gas chromatography mass spectrometry (GC-MS).

### Stool Collection and Processing

Stool samples were produced by subjects at home within 7 days of breath collection. Subject parents and guardians were provided a sterile, disposable spoon, collection tube (Meridian Bioscience, Para-Pak Cat# 23-290144) and toilet hat to complete collection and a styrofoam container with pre-chilled freezer packs to keep collected stool frozen. Stool samples were returned to the clinic within 24 hours of collection and then frozen at −20° C. Frozen stool samples in the clinic were transferred to the laboratory within 7 days and frozen at −80° C until processing.

At the time of processing, tubes containing subject stool samples were placed on dry ice. Stool from each individual fecal sample was placed in a sterilized mortar pre-chilled with liquid nitrogen and pulverized by mortar and pestle, adding additional liquid nitrogen to the mortar to prevent thawing. Pulverized human stool was then aliquoted into sterile 2 mL screw-cap tubes for downstream processes. All work on human stools was conducted within a biosafety cabinet by one of three individuals with previous experience pulverizing human stool (*28*). Aliquoted stool samples were stored at −80° C until further processing.

### Animal Subjects

Animal experiments were reviewed and approved by the Washington University Institutional Animal Care and Use Committee (Protocol #21-0394). Germ-free C57BL/6 mice were bred and maintained in-house in sterile, flexible vinyl isolators. Sterility was assured by monthly monitoring of mouse stool by confirming lack of amplification of the V4 region of the 16S rDNA gene, and aerobic and anaerobic culture. Mice were maintained on a diet of autoclaved mouse chow (LabDiet: Standard Diet 5021 – Autoclavable Mouse Breeder) and autoclaved water.

Conventional C57BL/6 mice were acquired at 8 weeks of age from either The Jackson Laboratory or Taconic Biosciences. Upon receipt, conventional mice were transferred into sterilized flexible vinyl isolators and maintained on the same autoclaved chow and water as germ-free mice for one week prior to the start of the experiment. Monocolonized gnotobiotic mice were maintained in a separate gnotobiotic facility using the Allentown Sentry Rack. These mice were maintained on a separate autoclavable mouse chow (LabDiet 5K67 – Autoclavable chow). Monocolonized mice were housed with no more than 5 mice per cage for 14 days.

Handling was performed in a sterile biosafety cabinet by trained personnel to maintain sterility of the mice and the interior of the cages. Mouse fecal pellets or cecal contents were collected from the lower colon by dissection into sterile 2 mL screw-cap tubes and flash-frozen in liquid nitrogen. Monocolonization at the time of harvest was confirmed by V4 16S rDNA sequencing of fecal pellets or cecal contents.

### Collection of mouse exhaled breath

Mice were anesthetized using an intraperitoneal injection of 300 µL of a cocktail containing 10 mg/mL ketamine and 1 mg/mL xylazine. Depth of anesthesia was assessed by regular assessment of the toe pinch response, and mice received additional anesthesia using an intraperitoneal injection of 100 µL of 10 mg/mL ketamine as necessary. Mouse vitals were monitored using the Kent Scientific PhysioSuite with a MouseStat heart rate monitor. Subject heart rate was sustained between 150 to 300 beats per minute during breath collection.

Breath was collected using the SciReq FX1 Flexivent murine ventilator. Prior to mouse breath collection, the sample path of the Flexivent was blocked and 3 volumes of 1.1L of clean (low VOC or scrubbed) air were collected through the instrument to serve as input air controls. To collect breath, anesthetized mice underwent surgery to place a 19G, 0.5 inch, blunt, metal intratracheal cannula before being connected to a Flexivent using established protocols (*74*). The input port of the Flexivent was then connected to a 25L SKC SamplePro Flexfilm bag (Cat# 237-25) filled with Zero Grade Air from AirGas (Cat# UZ300). The Flexivent was then operated under default settings (150 breaths per minute, 10 mL/kg tidal volume). After five minutes of venting the output line with subject breath, the output port of the Flexivent was connected to an empty 3L Flexfilm bag (Cat# 237-03). The time for collection was estimated for each mouse using the tidal volume with the set goal of acquiring 1.1 L of exhaled breath. The total time over which breath collection took place was between 36 and 66 minutes.

After breath collection was complete, each mouse was euthanized and fecal and cecal samples were harvested for further analysis. Flexfilm bags were set aside for 15 minutes to allow breath water vapor to condense before being drawn through CAMSCO inert stainless steel thermal desorption tubes (CAMSCO Cat# SIU60520-60-B) at a rate of 200 mL/min over five minutes, ultimately adsorbing volatiles from 1L of collected breath. TD tubes were stored at 4° C until shipped refrigerated for quantification. All breath samples were characterized within two weeks of collection, the prescribed limit of stability for breath samples on adsorbed TD tubes (*75*). The Flexivent and the connecting tubing was cleaned before and after each date of collection using a mild detergent (Simple Green Clean Finish disinfectant cleaner diluted 1:200 in ultrafiltered water) and ultrafiltered water.

### Isoprene dosing

Isoprene-treated mice received 1 mg of isoprene [Sigma-Aldrich, Cat# 59240-1ML-F] dissolved in 50 µL of vehicle-grade corn oil [Sigma-Aldrich, Cat# C8267-500ML] each day for 14 days intraperitoneally. Vehicle control mice simultaneously received 50 µL of vehicle-grade corn oil without isoprene. Mice were not dosed on the day of breath collection. On the day of harvest, breath was collected as previously described and full breath bags were used to load thermal desorption tubes.

### Bacterial Culture and Strains Human Clinical Isolates

*Akkermansia muciniphila* strain (AM02219), *Collinsella aerofaciens* strain (CA02219) *Ruminococcus torques* strain (RT04319), and *Bifidobacterium longum* strain (BL04319) were isolated from stool of human participants in the MARS clinical study (*28, 29*). AM02219 and CA02219 were isolated from the healthy participant MARS0022 and RT04319 and BL04319 were isolated from a participant with asthma, MARS0043. The methods for isolation are identical to the previously described approach used to isolate the *Bacteroides fragilis* strain BFM04319 described in Wilson et. al 2023. To summarize, a 10 mg/mL stock of homogenized stool in 10% glycerol was thawed in a Coy anaerobic chamber, plated on BHI/mucin agar containing 0.1% mucin, 0.1% resazurin, 0.05% L-cysteine hydrochloride and 1.2 mg/L histidine hematin and incubated at 37° C under strict anaerobic conditions for 6 days. Single colonies were isolated in BHI/mucin liquid media and stocked in 10% glycerol. Culture purity and identity was confirmed by V4 16S rDNA sequencing.

The *Escherichia coli* strain used here was a kanamycin-resistant derivative of a human cystitis isolate (UTI89attHK202::KanR) (*76*). The *Bacteroides thetaiotaomicron* strain used here (*B. theta* WT 5482 and tnaA KO) were generously provided through the Sperandio lab (*57*).

### V4 16S rDNA Sequencing of Bacterial Culture

DNA from cultured isolates was extracted from 1 mL of culture pelleted by centrifuging at 15,000 rpm for 1 minute. Media was drawn off and the pellet was washed twice in ultrafiltered water before the pellet was homogenized in phenol:chloroform:isoamyl alcohol by bead beating with 0.1 mm zirconium beads. The aqueous layer of bead-beat homogenate was isolated and purified using the 96-well QIAGEN PCR Clean up kit (Cat# 28181), quantitated by measuring absorbance at 260/280 nm, and normalized to 5 ng/µL. Normalized genomic DNA (10 ng) was PCR amplified using barcoded primers described in Hazan et. al (*77*). Amplified DNA was quantitated using the broad-range Quant-iT^TM^ dsDNA, pooled together, and purified twice using AMPure XP SPRI beads. Barcoded, amplified genomic DNA was sequenced using a MiSeq with 2×250 bp chemistry. All samples achieved a minimum read depth of 5000 reads. FastQ files representing V4 16S rDNA sequencing were demultiplexed and processed using DADA2 (*78*). Forward reads were trimmed to 200 base pairs, reverse reads were trimmed to 160 base pairs. Filtered and trimmed reads were binned into ASVs. Taxonomic classification was conducted using a custom reference database with a minimum bootstrap support of 80%. ASVs were normalized by total sum scaling.

### Collection of anaerobic culture headspace

Prior to culture, 250 mL Erlenmeyer flasks with magnetic stir bars were cleaned, autoclaved and allowed to degas in a Coy anaerobic chamber for a minimum of 48 hours. Anaerobic gas used for culture consisted of 75% nitrogen, 20% carbon dioxide, and 5% hydrogen. Bacterial strains were grown from a 10% glycerol frozen stock by thawing an aliquot in the anaerobic chamber and then diluting 100 µL of stock into 100 mL of BHI-mucin. Individual cultures were grown in sealed flasks for 48 hours at 37° C. Flasks were sealed with custom stoppers featuring a pair of stopcocks that could be manipulated to regulate airflow into and out of the flask. Multiple sterile media controls were processed and analyzed alongside bacterial culture.

At the time of collection, cultures were removed from the anaerobic chamber and incubated, stirring at low speed, at 37° C on a combination hotplate/magnetic stirrer for 15 minutes. For collection, one inlet of a stopcock was connected to an air pump that would draw air out of the flask and through a thermal desorption tube at a rate of 200 mL/min for 5 minutes. To ensure that the pump did not create a vacuum, the other inlet was connected to a Flexfilm bag containing a reservoir of anaerobic mixed gas drawn from the same tank that supported the anaerobic chamber. After collection was complete, the TD tubes were sealed and stored at 4° C until being shipped for analysis. All headspace samples were measured within 2 weeks of collection.

### Shotgun metagenomic sequencing of human and mouse fecal communities

Processing of human and murine stool was conducted as previously described (*28, 29*). Briefly, 50 - 100 mg of pulverized, frozen human stool or a frozen mouse fecal pellet were homogenized in phenol:chloroform by bead beating with sterile zirconium and steel beads. Crude extract of DNA was purified using the 96-well QIAGEN PCR Clean up kit (Cat# 28181) and quantitated by measuring absorbance at 260/280 nm using a Biotek Synergy H1 microplate reader.

Purified fecal DNA from each sample was normalized to 0.5 ng/μL. Fecal metagenomic adapter-ligated libraries were then generated by tagmentation using the Nextera Library Prep kit (Illumina, Cat# 121-1030/1031) with modifications to increase throughput (*79*). Libraries were purified using AMPure XP SPRI beads, quantitated using Quant-iT dsDNA assay High-sensitivity kit (Invitrogen Cat# Q33130), and then combined in an equimolar ratio. Library evenness was preliminarily assessed by sequencing on a MiSeq instrument with 2×150 bp chemistry and libraries were normalized and re-pooled by read count. Whole metagenomic sequencing and data demultiplexing of the libraries was conducted using an Illumina NovaSeq with 2×150 bp chemistry to achieve an average of 20 million reads per sample by the Genome Technology Access Center at the McDonnell Genome Institute (St. Louis, MO). All human stool samples and all mouse stool samples respectively were extracted, tagmented, and sequenced together to minimize batch effect.

### Processing and analysis of shotgun metagenomic sequencing of human and mouse fecal communities

Our analysis pipeline was based on the methods employed in a previous study (*28*). Briefly, demultiplexed FASTQ files were processed using kneaddata v0.10.0, trimmomatic v0.33 and bowtie2 v2.4.2 using default settings (*80, 81*). Read data from human stool samples was compared against the default human decoy genome to filter contaminant reads (*82*). FastQC (v0.11.7) was used to generate quality reports. After trimming and filtering, no samples had adaptor content, overrepresented sequences, or average Phred sequence quality score below Phred 24. Read data from mouse stool samples was compared against the default mouse C57BL database to filter contaminant reads. Genome coverage was estimated using nonpareil v3.4.1 (*83*). We estimated that the coverage achieved by each human stool sample after filtering ranged between 86.8% - 97.3%. Sequencing of the 36 colonized mice yielded a total 197 Gb of data after filtering, trimming, and decontamination using kneaddata. This represented a total of 784.1 million read pairs with a range of 11.1 to 147.1 million read pairs per fecal pellet. Using nonpareil, we estimated that samples achieved a median metagenomic coverage of 93.8% and a range of 88.8 to 99.5%. As with the human whole metagenome shotgun sequencing, we inferred the taxonomic composition of the gut metagenomes using metaphlan v3.1.0 (*84*) and inferred pathways and uniref90 gene clusters using humann3 v3.7 with the uniref90_201901b_full and 2023 Chocophlan databases (*84*). Reads filtered at each processing step and other summarized metrics are shown in **Table S2.**

### Human breath metabolite analysis

Human breath samples were run with a Quadrupole time of Flight Gas Chromatography Mass Spectrometry (GC-qTOFMS, Agilent Technologies, USA) attached to a Unity-xr (Markes international, UK). Before analysis, TD tubes were brought to room temperature and loaded to Unity-xr. A gaseous standard mixture (bromochloromethane, 1,4-difluorobenzene, chlorobenzene-D5) was added to each tube, followed by a dry purge purged for 5 min with N2 (Airgas) at 50 mL/min immediately prior to analysis. Tubes were desorbed at 270°C, 40 mL/min He flow, with recollection on a secondary Tenax cold trap at 10°C. Analytes were released from the secondary trap by heating to 295°C with 20% transferred to a gas chromatograph (7890B Series GC, Agilent Technologies, USA). The GC had a 30 m length x 0.25 mm ID x 0.25 µm film thickness DB-5MS column (Agilent, USA). The GC oven was programmed to hold at 35°C for 3 min, ramp 5°C/min to 175°C, then final ramp 60°C/min to 300°C, then hold at 300°C for 2 min.

The qTOF (Agilent Technologies, USA) used an electron ionization source set at a temperature of 230°C. The quadrupole was set to a temperature of 150°C and the collision cell had a nitrogen flow of 1.5 mL min^-1^. The emission current was fixed at 35 µA and the electron energy at 70eV. The TOFMS had a sampling frequency of 50 Hz and a mass recording range of 35 - 400 amu. The abundances of isoprene in each sample were calculated by the integrating the respective base ion peaks at the proper retention time (determined by comparison to a true standard) using the Extract Ion Chromatogram feature for ions 67, 68 with Mass Hunter Qualitative analysis software (version B.07.00).

The raw data generated were first converted to an open standard format such as mzdata.xml. Thereafter, the search for compounds in the breath samples was carried out by the ion peaks detection technique using the XCMS (*85*) module of the free statistical software R. The three main steps conducted with XCMS on our data were: peak detection, retention time alignment, and peak matching. Denoising (smoothing) and baseline correction were first performed to optimize peak detection. Thereafter, peak detection (step 1), was performed using the matchedFilter algorithm to extract ion signals. This step was optimized to determine the largest number of peaks in each sample without incorrectly duplicating peaks and with the separation of overlapping peaks. Next, retention time alignment (step 2) was performed using the retcor method. This step was required to ensure that the retention times were aligned across all samples because an intragroup variation in peak retention time was observed. Finally, peaks previously detected in each sample were grouped across all the samples through peak matching (step 3). Thus, peaks with similar characteristics (same m/z and similar retention time) were considered to be the same feature. A data matrix with the intensities of the features, m/z and retention times in each sample was obtained after performing steps 2 and 3. Following the data pre-processing steps, ions corresponding to contaminants were removed which resulted in a total of 2734 features for further analysis.

### Mouse breath and bacterial headspace metabolite analysis

#### Bacterial headspace analysis

(method used in Fig. 5C). All TD sorbent tubes were auto-processed using a Turbomatrix 650 thermal desorption apparatus (PerkinElmer, Waltham, MA) unit interfaced to a Leco Pegasus 4D GC × GC-TOFMS (time-of-flight mass spectrometer; Leco Corporation, St. Joseph, MI) used in 1-D GC mode (i.e., without application of a secondary column). Before analysis, TD tubes were brought to room temperature. A gaseous standard mixture (fluorobenzene, toluene-d8, 4-bromoflurobenzene and 1,2-dichlorobenzene-d4) was added to each tube by the thermal desorber immediately prior to analysis. Each tube was then purged for 5min with He at 30 mL/min prior to thermal desorption. The tube was desorbed (270 °C, 10 min) with 40mL/min of helium and re-concentrated on a focusing trap (Tenax® TA, 10 °C) within the Turbomatrix 650. The focus trap was then thermally desorbed at 295 °C for 7 min at 18.6 psi He pressure with a split flow of 8 mL/min. The effluent passed through a heated fused silica transfer line (225 °C), which is directly connected to the GC column by a press tight connector (Restek, Bellefonte, PA). The GC oven temperature for each sample was programed as follows: held at the initial 40°C for 5 min, then increased by 12°C/min increment to reach 220°C, followed by 20°C/min increment to reach and hold at 280°C for 4 min, then 20 ⁰C/min to 220 ⁰C and hold at 220 ⁰C for 1 min. The GC column (Rxi-624 SilMS, 30m x 0.25mm id x 1.4µm, Restek, Bellefonte, PA) interfaced directly into the MS transfer line (235°C). The MS detector was at 1680 V. The data was recorded at 50 spectra/s with a mass recording range 34–400 amu. The raw data was then exported to CDF format.

The CDF data generated was processed using the XCMS in the same way as for human samples.

#### Mouse breath and bacterial headspace analysis

(method used in Figures 4, 5B, 6, 7 and Fig. S3). Prior to analysis, sorbent tubes were brought to room temperature and loaded into autosampler (Utra-xr, Markes International, UK). A gaseous standard mixture (1.01 ppm bromochloromethane, 1.04 ppm 1,4-difluorobenzene, 1.04 ppm chlorobenzene-D5, 0.96 ppm 4-bromofluorobenzene) was immediately added to each tube, followed by a purge pre-desorption step consisting of 10 min with N_2_ at 50 mL/min, to remove water content in breath samples. Tubes were thermally desorbed for 10 min at 270°C (Unity-xr, Markes International, UK) and transferred to a general purpose focusing trap that matched the sorbent of the sample tube, held at 10°C and subsequently heated to 300°C, to minimize band broadening. The split flow after the cold trap was 4.8 mL/min.

Analysis by two-dimensional gas chromatography (GCxGC) was conducted using an Agilent 7890B GC system, fitted with a flow modulator and a three-way splitter plate coupled to a flame ionization detector and a time-of-flight mass spectrometer with electron ionization (SepSolve, UK). Chromatographic analysis was performed using a Stabilwax (30 m × 250 μm ID × 0.25 μm df) as the first dimension (1D)-GC column and a Rtx-200 MS (5 m × 250 μm ID × 0.1 μm df) as second dimension (2D)-GC column, both purchased from Restek (Bellefonte, PA, US). The following GC oven temperature program was used: initial temperature 40°C was ramped to 215°C at 3°C/min and held for 1 min. The oven temperature was then increased to 260°C at 50°C/min then held for 10 min. The total run time for the analysis was 75 min. Helium carrier gas was flowed at a rate of 1.2 mL/min. The flow modulator (Insight, SepSolve Analytical, UK) had a loop with dimensions 0.53 mm i.d. x 110 mm length (loop volume: 25 µL), and the modulation time was 2 s.

The GCxGC was interfaced with a BenchTOF-select time-of-flight mass spectrometer (SepSolve Analytical, UK). The acquisition speed was 50 Hz and mass range was 35-350 m/z. The ion source and transfer line were set at 250 °C and 270 °C respectively and filament voltage at 1.7 V. Electron ionization energy was 70 eV. ChromSpace (SepSolve Analytical, UK) was used to synchronize and control the INSIGHT modulator, thermal desorption, GC, and TOF.

### Untargeted GCxGC-MS data analysis

GCxGC–MS deriving chromatograms were first aligned to one user-selected reference chromatogram in ChromCompare+ software version 2.1.4 (SepSolve Analytical Ltd, UK) based on the 1D and 2D retention times and the available spectral information. The alignment algorithm was used to overcome retention time drift observed across the dataset.

A tile-based approach was applied to the aligned chromatograms to enable the raw data to be imported into the chemometrics platform directly, without the application of any pre-processing methods, such as integration and identification. The tile size was 32 s in 1D and 0.5 s in 2D with 25% overlap. The signal for every individual m/z channel of each tile was integrated for comparison across every chromatogram in the dataset. This process generated a list of features labeled according to the tile retention times and m/z channel.

The data matrix was cleaned of possible artifacts and siloxane derived from the sorbent and filtered so that individual m/z channels with < 30000 intensity were removed. A feature was retained if it was present in more than 50% of the samples in either group (e.g. media vs culture) and data were then normalized using internal standard (4-bromofluorobenzene).

Data reduction was performed using the proprietary feature selection algorithm in ChromCompare+ software. The algorithm uses a multivariate method to consider the covariance between features (*86*); this approach retained the most significant features of the known sample classes. Unique peaks were putatively identified via comparison with the NIST20 library based on the combination of the mass spectra similarity match ≥60% and further identity confirmation was obtained with pure standards.

Random forests were produced using the randomForest package v 4.7.1.1 in R v4.3.2. Feature selection was performed using Boruta (v8.0.0) (*87*) over 100 iterations. Random forests used for feature selection used 1000 trees and an mtry of 3 and the final model used 1000 trees with an mtry of 3. The random forest with selected features for each VOC was performed 20 times to estimate the variance explained (R^2^-statistic) with confidence intervals. Shapley additive explanations from the random forests were estimated using the treeshap package (v0.3.1) on the models after feature selection (*88*). The importance of each feature to a model was estimated as the average absolute value of the SHAP scores for that feature.

### Statistics

Statistics and all data analysis were performed in R version 4.3.2. Data are represented as means with standard deviation unless otherwise specified. Multiple hypothesis correction was performed by the Benjamini-Hochberg method when necessary. Violin and box plots display the IQR and whiskers display 1.5* the IQR. The following symbols are used to denote significance: * p<0.05, ** p<0.01, *** p< 0.001, **** p< 0.0001. Principal coordinates analysis data employed UniFrac distances (*44*) for taxonomic metagenomic data and Manhattan distances for pathway and Uniref90 metagenomic data. Manhattan distances for principal coordinates analysis of breath data. PERMANOVA were performed using the adonis2 function and PROCRUSTES analyses were performed using the protest function in vegan (*89*) v2.6.4. Age and BMI were treated as continuous features whereas all others were categorical. BMI was normalized to percentile based on age and sex of patient.

## LIST OF SUPPLEMENTARY MATERIALS

**Table S1. Summarized demographic data for MACK subjects**

**Table S2. Summarized metrics for shotgun metagenomic sequencing of human and mouse fecal communities**

**Table S3. List of VOCs known to be found in breath and analyzed in this study**

**Table S4. List of VOCs found in monocolonization and anaerobic culture headspace experiments**

## Data Availability

Code base and computational notebooks for the analysis of all data and production of all figures is available through Zenodo (DOI: 10.5281/zenodo.12702321). All sequencing data associated with this study are available from the European Nucleotide Archive under accession number PRJEB76956. Converted GC-MS files (mzML format) are available through the MassIVE repository (https://massive.ucsd.edu/ProteoSAFe/static/massive.jsp) under the following identifiers: MSV000095340 (breath chromatograms from healthy children) and MSV000095341(chromatograms from bacterial headspace metabolites)

## ACKNOWLEDGEMENTS

We would like to thank Chad Schaber for helpful discussions; Vanessa Sperandio for providing us with the tnaA deficient *B. thetaiotaomicron*; and the Baldridge lab for assisting us with bacterial culture. We also thank Jessica Hoisington-Lopez in the DNA Sequencing Innovation Lab at the Center for Genome Sciences and Systems Biology and Genome Technology Access Center at McDonnell Genome Institute for assistance with sequencing. We are grateful for the assistance of Tarisa Mantia and Caitlin O’Shaughnessy in conducting the MACK study and Mike White of the gnotobiotic core.

## Author contributions

A.J.H., A.Z.B., A.L.K. and A.R.O.J. conceived the original concept and initiated this project. A.J.H., A.L.R., M.A.L., R.T.M., N.J., J.S.B., and C.P.T. developed the technique for collecting breath from mice and assisted in breath collection from mice. A.J.H. performed analysis on metagenomic sequencing data. A.L.R., A.J.H., and A.Z.B. cultured and collected bacterial headspace volatiles. W.L., A.Z.B. and Y.L. collected and analyzed GC-MS data. A.J.H., S.A.W., and A.L.R. visualized the data. A.J.H., A.Z.B., S.A.W., A.R.O.J., and A.L.K. wrote the manuscript. All authors reviewed the final manuscript.

---

**Fig. S1:**
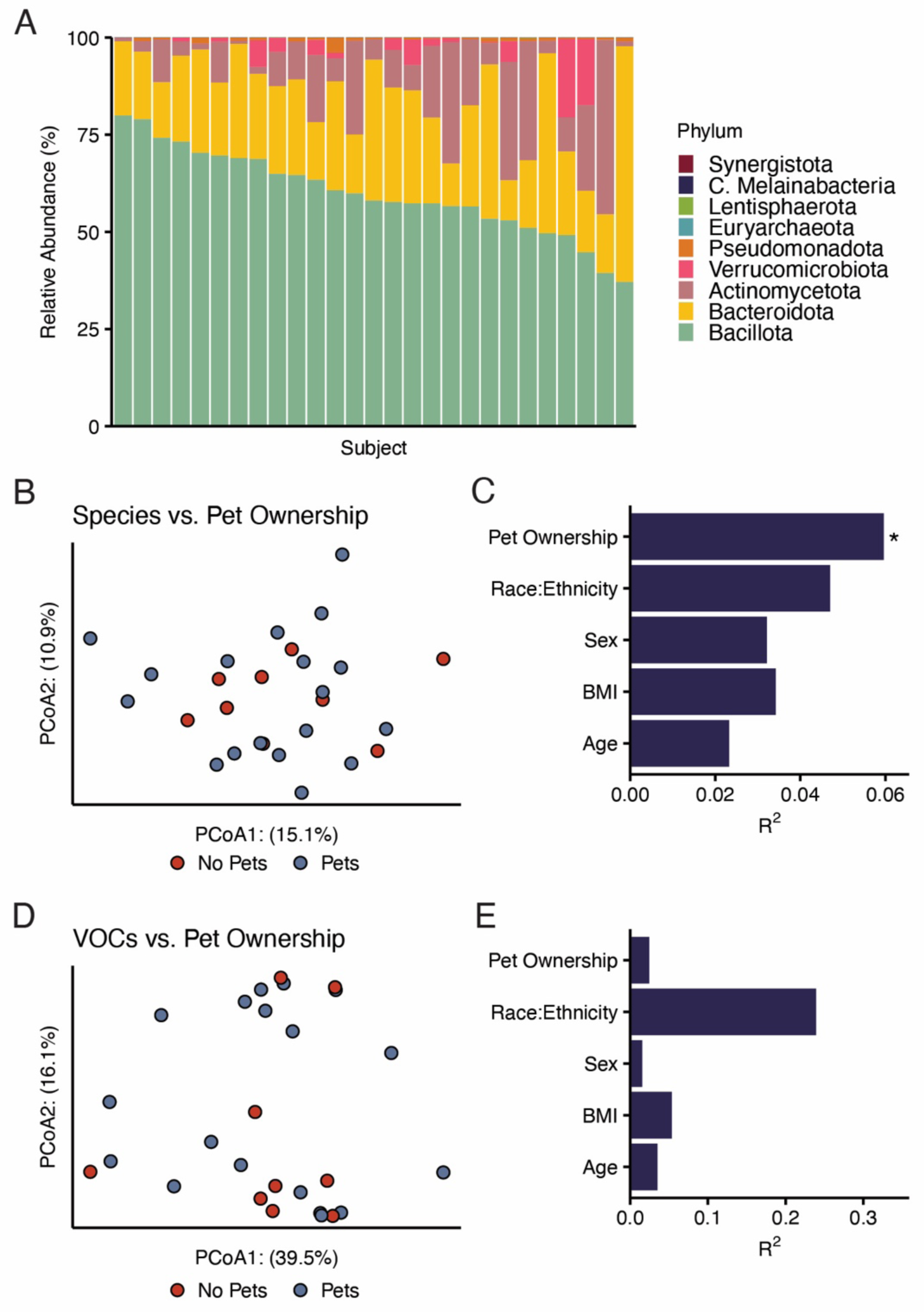
Overview of the gut metagenomes of 27 healthy pediatric subjects. A) Taxonomic relative abundance mosaic plot demonstrating the compositions of the gut microbiota of individual subjects as predicted by metaphlan3. B) Principal coordinates analysis on the UniFrac distances between subject gut taxonomic compositions, specifically highlighting pet ownership, which predicts an unexpected degree of variance in subject gut composition. C) Results of a Type III PERMANOVA comparing subject demographic features against the taxonomic composition of subject gut microbiota. D) Principal coordinates analysis on the Manhattan distances between subject breath volatilomes, specifically highlighting pet ownership. E) Results of a Type III PERMANOVA comparing subject demographic features against the Manhattan distances between subject breath volatilomes.

**Fig. S2:**
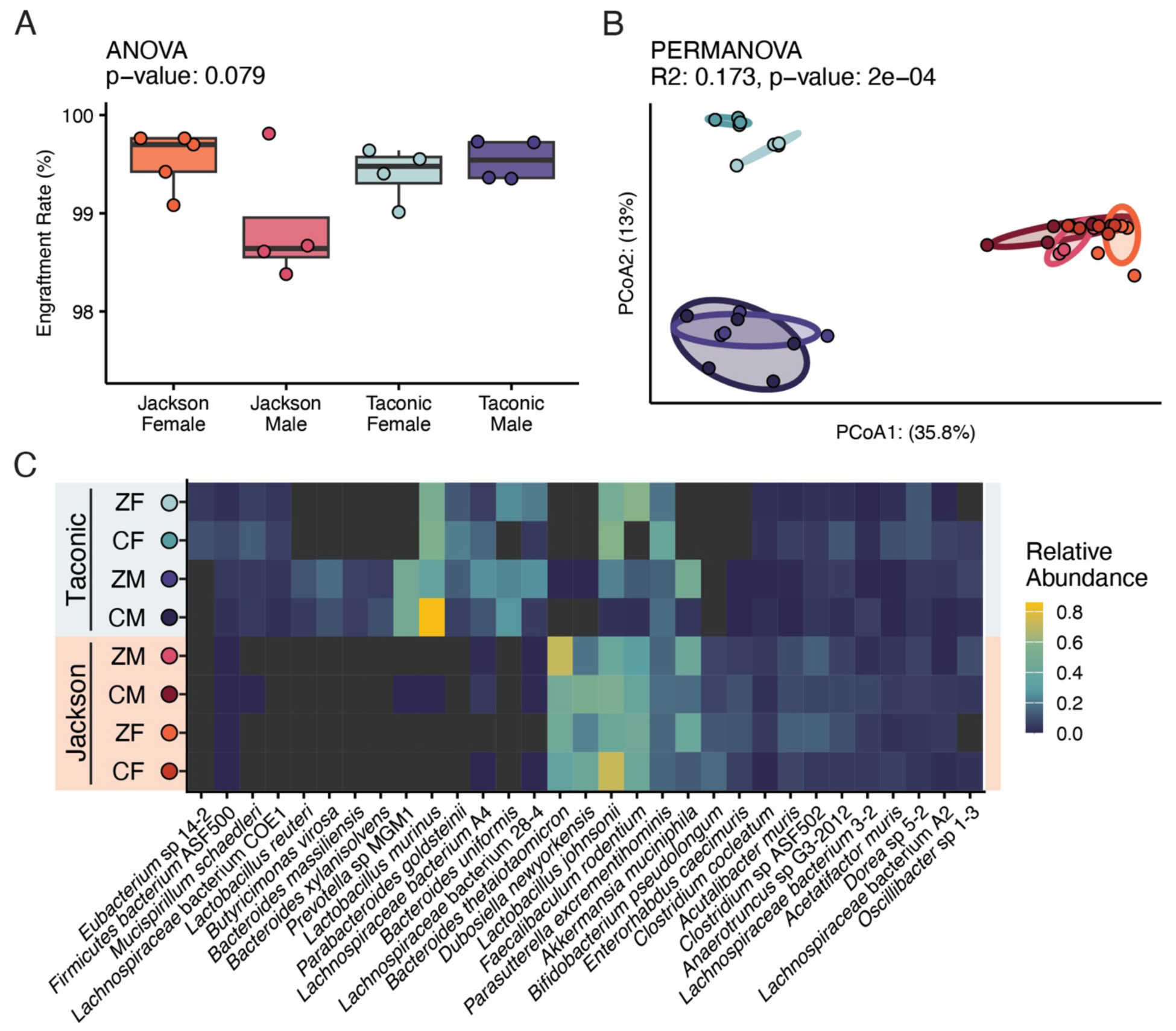
Overview of the metagenomic profile of conventionally raised mice and conventionalized gnotobiotic mice. A) Boxplot comparing engraftment rate of donor microbiota against conventionalized mouse microbiota. B) Principal coordinates analysis visualizing UniFrac distances between all colonized mice. C) Heatmap depicting the relative abundances of all colonized mice averaged by microbiota group.

**Fig. S3:**
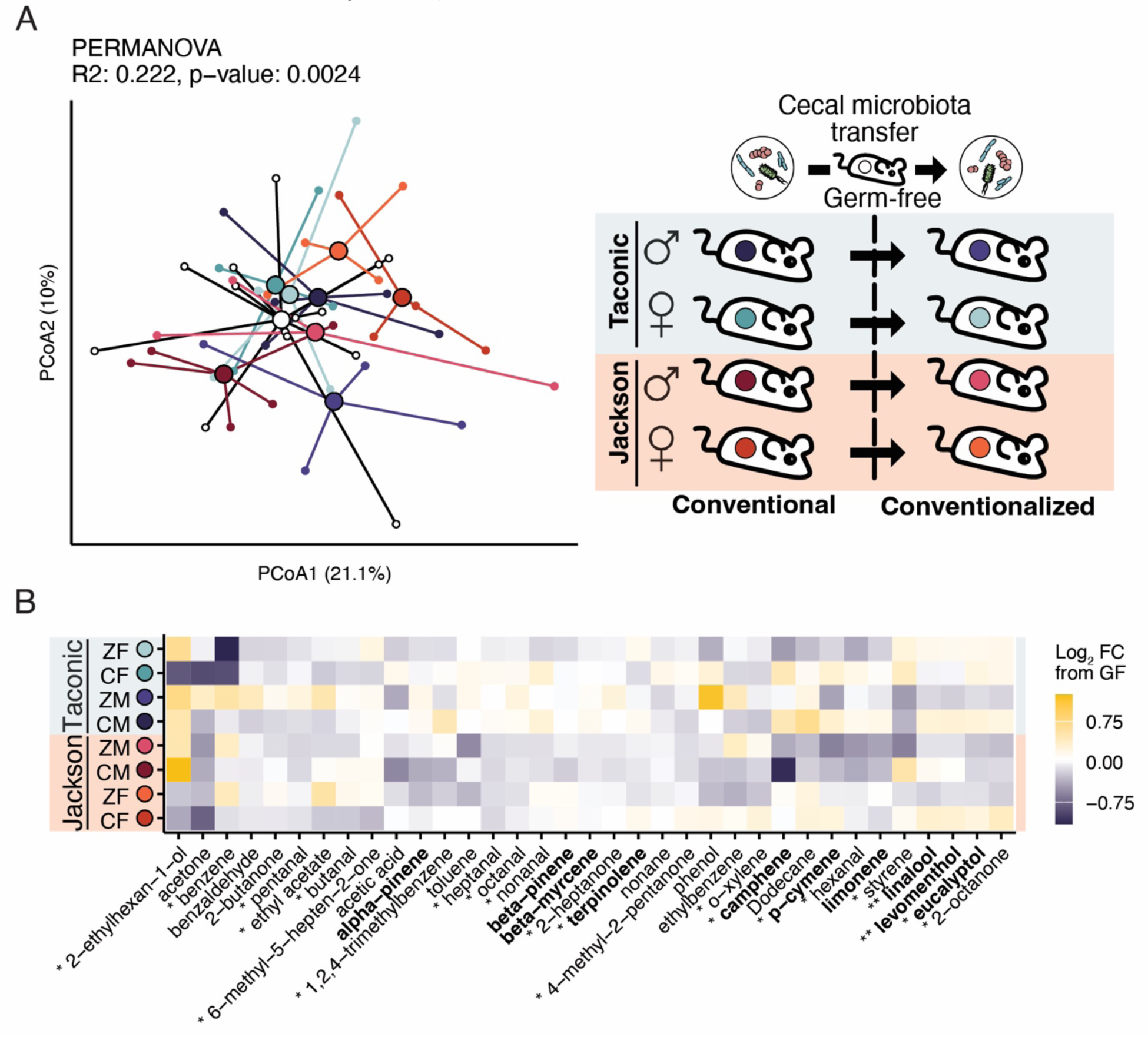
Overview of the breath volatilome profile of conventionally raised mice and conventionalized gnotobiotic mice. A) Principal coordinates analyses visualizing Manhattan distances between the breath volatilomes of conventionally-raised and gnotobiotic mice. Germ-free mouse breath is represented in white in A and the centroid in black in B. B) Heatmap visualizing the average Log2 fold difference between a group of colonized mice and germ-free mice for individual volatiles in exhaled breath. Tests between groups were conducted using Kruskal-Wallis with Benjamini-Hochberg multiple hypothesis correction. Terpenes are bolded. VOC comparisons yielding adjusted p-values below 0.05 are noted with an asterisk*.

